# Maternal Stress-Related Neural Reactivity and Caregiving Sensitivity in Early Childhood: Associations with Peri-Pubertal Disruptive Behaviors

**DOI:** 10.64898/2025.12.02.25341440

**Authors:** Ryan J. Murray, Setareh Ranjbar, Shannen Graf, Sandra Rusconi Serpa, Camille Piguet, Sébastien Urben, Daniel S Schechter

## Abstract

Disruptive behavior disorders among youth are a pressing public-health concern and a key pathway of intergenerational risk. In the 8-year longitudinal Geneva Early Childhood Stress Study, we tested whether maternal psychological, behavioral, and neurobiological factors assessed in toddlerhood predicted peri-pubertal disruptive behaviors. Mother–child dyads (n = 28; child age at Phase 1 = 12–44 months; Phase 3 = 9–15 years) completed clinical interviews; mothers underwent fMRI while viewing emotion-related (prosocial/romantic and threatening vs neutral) and threat-related (threatening vs prosocial/romantic) adult interactions; maternal sensitivity (MS) was coded from filmed play; child disruptive symptoms were assessed with the Kiddie Schedule for Affective Disorders and Schizophrenia (K-SADS) at follow-up. Ten a priori neural clusters were reduced via principal components analysis, yielding a theory-consistent Neural-Threat component and a Neural-Preoccupation component reflecting precuneus activity. Using exploratory forward stepwise regression adjusted for child age, sex, and socioeconomic status, Neural-Threat and MS each uniquely predicted fewer disruptive behaviors (protective), whereas Neural-Preoccupation predicted more symptoms (risk). A planned moderation test showed that MS buffered the Neural-Threat–behavior association, with stronger protection at higher sensitivity. Maternal posttraumatic stress severity in early childhood showed no direct association with later disruptive behaviors. These findings suggest that stress-responsive maternal brain function relates to children’s disruptive outcomes and that sensitive caregiving may potentiate protective neural processes. Results are preliminary and warrant replication in larger, more representative samples, but they highlight neurobehavioral targets for early, trauma-informed prevention, i.e., enhancing maternal sensitivity and supporting adaptive threat monitoring in mothers.

## 1. Introduction

Youth violence is an escalating global public health concern. Whereas, converging evidence in the United Kingdom alone show increasing youth-related violence and weapon-related injuries (Gwata et al., 2024), an estimated 193,000 youth homicides (15–29 years) are now recorded globally each year; homicide now ranks among youths’ leading causes of death (World Health Organization, 2024). This is consistent with Swiss rates. Indeed, the number of serious acts of violence in Switzerland increased by 16.6% to 1,942 offences in 2022, the highest value since such statistics started to be recorded in 2009 (SWI swissinfo.ch, 2023). Because aggressive trajectories typically become clearer beginning in the peri-pubertal years, it is essential to prioritize early prevention of disruptive behavior disorders (DBD) (i.e., a spectrum of problematic behaviors marked by chronic impulsivity, rule-breaking, and defiance (American Psychiatric, 2013), in order to curb later more serious violence.

Disruptive behavior pathways are also central to intergenerational trauma research. They forecast serious adult violence, criminality, and revictimization (Moffitt, 2018; Tremblay et al., 2004) and are over-represented among children of parents exposed to early (i.e., childhood) interpersonal violence (IPV) (Racine et al., 2023). Exposure to IPV is a global epidemic, particularly in women. A review of 366 studies (N > 2 million women from 161 countries) reported IPV exposure of up to 27% in women and implicating intimate partner violence in nearly half of female homicides (Gilbert et al., 2009; Kourti et al., 2023; Michl-Petzing et al., 2019; Stith et al., 2009; World Health Organization, 2010, 2014). Understanding how maternal IPV-related trauma shapes peri-pubertal disruptive behaviors is therefore a public-health priority and a key to prevention.

Parents who were themselves exposed to IPV in childhood (maltreatment or witnessing domestic violence) are at elevated risk for PTSS in adulthood. (Teicher & Samson, 2013); IPV-related PTSS increases parenting distress and disrupts sensitive caregiving during formative developmental windows, thereby heightening offspring risk for psychopathology through both direct and indirect routes. Directly, parents abused in childhood are nearly three times more likely to expose their own children to violence (Assink et al., 2018; Bartlett et al., 2017; Oliver, 1993; Yang et al., 2018). In a South-Australian cohort (N=38’556), offspring of mothers with verified maltreatment and out-of-home placement were 6 times more likely to be maltreated themselves (Armfield et al., 2021). Over 36% of women who had child protective records during their own childhoods had children who also had child protective services involvement; among those children, 41–75% (thus 15-27% of the entire sample) of their cases were verified for exposure to maltreatment and/or family violence, suggesting that 15-27% of all children may ultimately experience intergenerational trauma directly (Armfield et al., 2021).

Indirectly, maternal IPV-PTSS and comorbid major depressive disorder symptoms (MDDS) augment risk even when children are not directly exposed to violence. Such maternal psychopathology is linked to elevated internalizing and disruptive behaviors in youth (Fenerci & Deprince, 2018; Glaus et al., 2021), independent of socioeconomic status (Sierau et al., 2020). Consistent evidence ties child outcomes to maternal PTSS (Van Ee et al., 2016), MDDS (O’Connor et al., 2017), to heightened parenting stress (Samuelson et al., 2017) and impoverished caregiving (e.g., low sensitivity [Moser et al., 2023]). And so, by affecting parenting and thus the quality of the parent-child relationship, collectively, these findings underscore the importance of addressing indirect parental psychopathology-mediated processes in the wake of parental childhood and adult exposure to IPV.

Maternal sensitivity, a core component of caregiving behaviors, has been consistently shown to be a bulwark against the indirect effects of maternal IPV exposure (Gustafsson et al., 2015; Hibel et al., 2011; Manning et al., 2014), with high maternal sensitivity potentially buffering the risk of maternal IPV-related effects on child behavior (Manning et al., 2014). Defined by Ainsworth as the accurate and developmentally appropriate response to infant cues (Ainsworth, 1978), maternal sensitivity is often impaired in IPV-affected mothers, who show more withdrawn or intrusive behaviors (Moser et al., 2015), misread child emotional signals, and make appraisal errors. Such impoverished caregiving has been linked to later child psychopathology (Moser et al., 2023; Pointet Perizzolo et al., 2022; Suardi et al., 2020).

Maternal neurobiological reactivity to stress is a largely overlooked mechanism in the intergenerational transmission of trauma, however, particularly for child disruptive behaviors that forecast later IPV (Schechter & Rusconi, 2011). While maternal PTSS (with or without MDDS) reliably mediates trauma-related risk for children’s internalizing problems (Choi et al., 2019; Giallo et al., 2020; Morelen et al., 2016; Sierau et al., 2020), comparable mediation is rarely observed for disruptive behaviors. This difference suggests latent drivers, such as maternal stress-related brain function, may better predict the impulsive, rule-breaking, and aggressive patterns that escalate to serious violence (Bachem et al., 2024).

Integrative work now calls for models that embed neurobiological markers alongside psychological and caregiving factors (Pereira et al., 2021; Schechter et al., 2011). For instance, maternal sensitivity correlates with fronto-limbic activation to emotional cues (Moser et al., 2015) and can buffer children from the behavioral impact of maternal trauma, yet PTSS-related hypo- or hyper-reactivity in these circuits may erode that protection. Whether stress-responsive neural activity, as a biological correlate of IPV-related symptoms, interacts with maternal sensitivity to shape offspring behavior, however, remains untested.

### 1.1. The Current Study

The present paper, based on an 8-year longitudinal study, asks whether mothers’ stress-related neural responses in toddlerhood forecast their children’s peri-pubertal disruptive behaviors and whether this link depends on maternal sensitivity. By testing both the direct predictive value of maternal brain reactivity and its interaction with sensitive caregiving, we aim to clarify how neurobiological and behavioral processes jointly shape intergenerational risk for impulsive, defiant, or aggressive behaviors.

By focusing on disruptive behaviors, our study addresses a behavioral outcome that (a) imposes heavy societal and economic burdens (World Health Organization, 2024), (b) has clear ties to intergenerational cycles of family violence (Assink et al., 2018; Bartlett et al., 2017; Oliver, 1993; Yang et al., 2018), and (c) may be particularly sensitive to early variations in maternal threat processing and caregiving style. Clarifying these neurobehavioral pathways is therefore essential for designing trauma-informed early interventions that can break the cascade from early adversity to adolescent behavioral dysregulation.

In this current study, we adopted a primary exploratory, data-driven model-selection approach to identify which maternal factors best account for variance in peri-pubertal disruptive behaviors. Specifically, we examined (i) whether maternal trauma-related psychopathology (IPV-PTSS) shows a positive association with child disruptive behaviors (e.g., emotional/behavioral dysregulation (Van Ee et al., 2016); (ii) whether maternal stress-related neural reactivity in toddlerhood (e.g., prefrontal, anterior cingulate systems) contributes unique variance to peripubertal disruptive behaviors (Schechter & Rusconi, 2011) and (iii) whether maternal sensitivity moderates any neural–behavior link—such that higher maternal sensitivity might buffer adverse associations with threat-related reactivity.

## 2. Methods

### 2.1. Participants

The present study is a two-phase investigation, nested within the larger Geneva Early Childhood Stress Project (Moser et al., 2019; Schechter & Rusconi, 2014) which was a three-phase prospective, longitudinal study comprising 113 mother–child dyads at baseline (Phase 1). Data analyzed here were drawn from two time points (Phases 1 and 3), and all participating families provided informed consent and received compensation in the form of money and/or age-appropriate toys and books.

At Phase 1 (P1), conducted between 2010 and 2014, mother–child dyads were recruited from the Geneva community through flyers distributed in pediatric clinics, maternity wards, and public spaces. Inclusion criteria required that participants be biological mothers living with their child, who was between 12 and 44 months of age. Mothers had to be fluent in French or English, with no history of psychosis, substance abuse, or significant physical or intellectual impairments.

At Phase 1 (P1), 63 of the initial 113 mothers underwent MRI scanning; nine were excluded due to poor scan quality (e.g., excessive motion, drowsiness). Of the remaining 54 dyads with valid MRI data, 48 returned for the Phase 3 (P3) follow-up. Twenty of these 48 dyads were subsequently excluded from the present analysis due to extensive missing exposure data (>40% [Graham, 2009]) either at P1 (16 dyads) or at P3 (4 dyads). Ultimately, complete and valid datasets from both P1 and P3 were available for 28 dyads (11 girls; child mean age at P3=10.86 ± 1.56 years, range: 9-15 years; see Figure 1).

**Figure 1.**
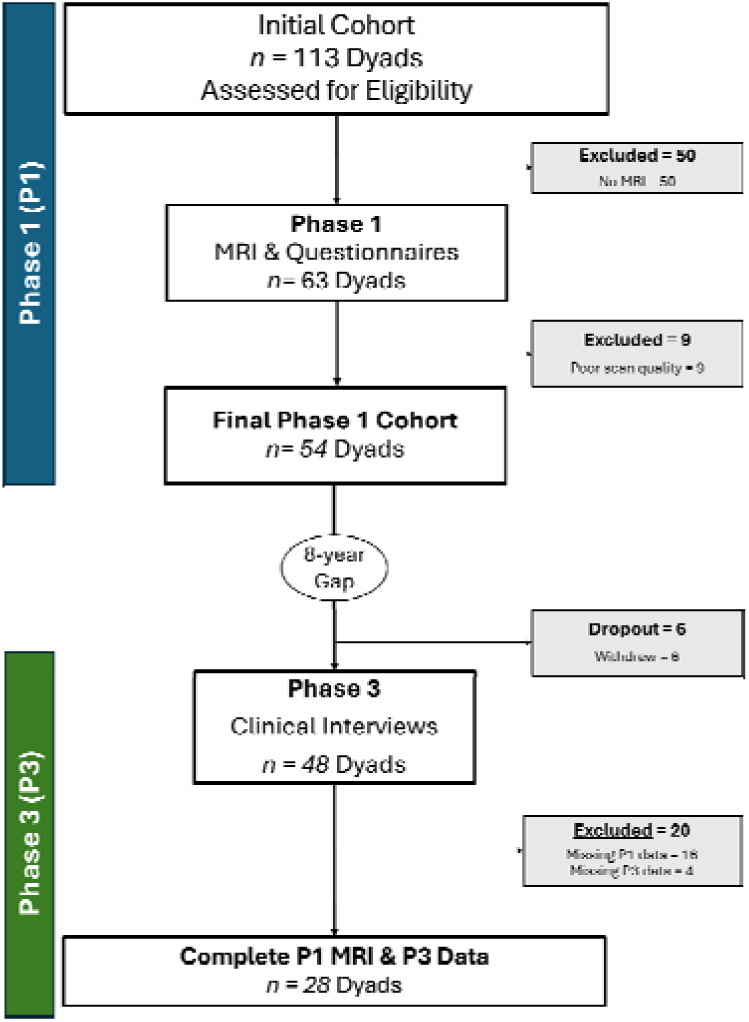
Two-Phase Participant retention flowchart.

### 2.2. Procedure

At P1, dyads participated in a comprehensive, multi-method assessment protocol within one month. Mothers underwent functional MRI scanning while viewing film clips depicting emotion-laden (escalating interpersonal conflict and romantic) vs neutral (no emotionally charged behaviors) adult male-female interactions and completed a battery of self- and clinician-administered instruments assessing trauma history, psychopathology, and parenting-related variables. Children were evaluated through video-recorded mother–child interaction tasks, clinical interviews, and behavioral observations conducted by trained researchers.

At P3, peripubertal disruptive behaviors was measured, via the semi-structured Schedule for Affective Disorders and Schizophrenia for School Aged Children (6-18 Years), lifetime version (K-SADS-PL [Kaufman et al., 1997]).

All procedures were approved by the Institutional Ethics Committee of the Geneva University Hospitals (protocol #14-271) and conducted in accordance with the Declaration of Helsinki (World Medical Association, 1999).

### 2.3. MRI

A detailed description of the MRI procedure, acquisition and preprocessing parameters in P1 has been provided in earlier papers (Moser et al., 2015) with complementary information in the Supplementary Materials. Mothers first underwent an hour-long screening and two 2-hour visits, which included clinical interviews and a behavioral protocol involving mothers and children.

In the MRI machine, participants viewed 23 clips of emotionally charged male–female interactions (cf., Moser et al., 2015) in pseudorandomized order. Details on image acquisition and preprocessing are available in supplementary data. Twenty-three silent 20-second video excerpts featuring male–female interactions were extracted from films and categorized as threatening (8 clips), prosocial/romantic (8 clips), or neutral (7 clips) based on an unpublished rating study detailed in the supplementary data. Neural reactivity to emotional and neutral stimuli was categorized into two contrasts: (i) Emotion-specific (threatening/prosocial/romantic>neutral); (ii) Threat-specific (threatening>prosocial/romantic) (Moser et al., 2015).

To assess the influence of maternal neurobiological functioning, we analyzed beta values from 10 clusters retrieved from emotion- and threat-specific neural activity from published studies (Table 1) (Moser et al., 2015; Moser et al., 2014).

**Table 1.**
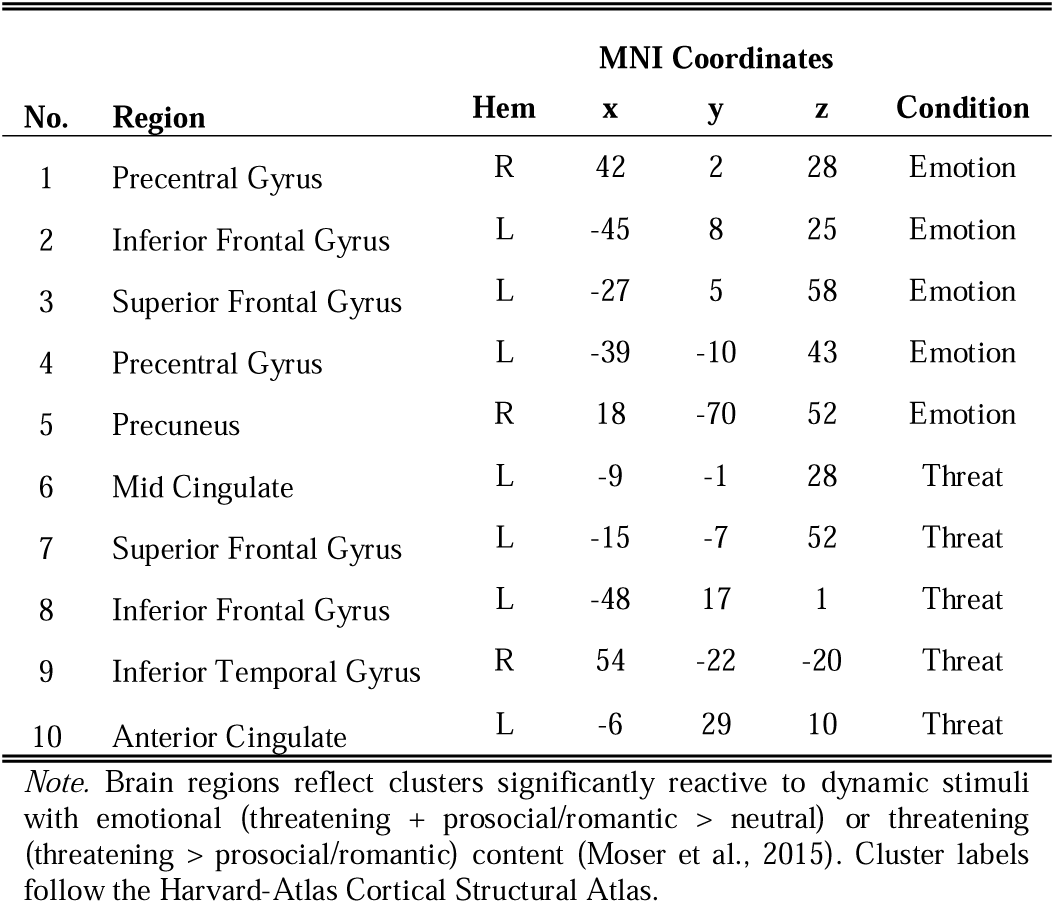
Emotion- and Threat-Related P1 Maternal Brain Clusters.

| No. | Region | Hem | MNI Coordinates |  |  | Condition |
| --- | --- | --- | --- | --- | --- | --- |
|  |  |  | x | y | z |  |
| 1 | Precentral Gyrus | R | 42 | 2 | 28 | Emotion |
| 2 | Inferior Frontal Gyrus | L | -45 | 8 | 25 | Emotion |
| 3 | Superior Frontal Gyrus | L | -27 | 5 | 58 | Emotion |
| 4 | Precentral Gyrus | L | -39 | -10 | 43 | Emotion |
| 5 | Precuneus | R | 18 | -70 | 52 | Emotion |
| 6 | Mid Cingulate | L | -9 | -1 | 28 | Threat |
| 7 | Superior Frontal Gyrus | L | -15 | -7 | 52 | Threat |
| 8 | Inferior Frontal Gyrus | L | -48 | 17 | 1 | Threat |
| 9 | Inferior Temporal Gyrus | R | 54 | -22 | -20 | Threat |
| 10 | Anterior Cingulate | L | -6 | 29 | 10 | Threat |

*Note.* Brain regions reflect clusters significantly reactive to dynamic stimuli with emotional (threatening + prosocial/romantic > neutral) or threatening (threatening > prosocial/romantic) content (Moser et al., 2015). Cluster labels follow the Harvard-Atlas Cortical Structural Atlas.

### 2.4. Variables and Measures

#### 2.4.1. Predictor Variables

Tested predictor variables, described below, were PTSS severity, Parental Distress, Maternal Sensitivity, and stress-related neural activity, all measured during P1.

##### PTSS Severity

Lifetime symptoms of IPV-related posttraumatic stress disorder (IPV-PTSS) of the mother was assessed at P1 via the clinician-administered PTSD Scale (CAPS) (Pupo et al., 2011; Weathers et al., 2001) in accordance with DSM-IV-TR criteria (Guze, 1995). Current PTSS symptoms were assessed via the Post-traumatic Symptoms Checklist-Short version PCL-S (Lima et al., 2012). Only women whose PTSS related to interpersonal violence were included in the study. IPV-PTSS was diagnosed if CAPS score was ≥55 and PCL-S score was ≥40. Absence of clinical-level PTSS (i.e., Non-IPV-PTSS) was determined if CAPS score was <30 and PCL-S score was <25. Scores between these ranges, although subthreshold IPV-PTSS, were included in the IPV-PTSS group. Group differences were only measured to assess baseline sample characteristics with regards to predictor variables and covariates.

##### Parental Distress

Parenting distress at P1 was assessed using the self-report Parental Distress subscale of the Parenting Stress Index–Short Form (PSI-SF) (Abidin, 1995). While the full PSI-SF includes three dimensions of parenting stress, Parental Distress was selected for its conceptual relevance to this study’s focus on maternal psychopathology and emotion regulation. Specifically, this subscale captures the intrapsychic experience of distress related to the parental role, such as role strain, anhedonia, and subjective emotional burden, all of which are particularly salient in IPV-exposed populations.

##### Maternal Sensitivity

During P1, maternal sensitivity was assessed using the CARE-Index, a validated observational system coding caregiver behavior, during a 5-minute mother–child free-play task (Crittenden, 2006). Mothers played with standard toys while being video-recorded prior to MRI scanning. Two trained, independent raters, blind to group status, coded videos using standardized procedures. Inter-rater reliability for sensitivity was high (ICC=0.86; *r*=.925, *p*<.001) (Moser et al., 2023). The CARE-Index provides scores across sensitivity, control, and unresponsiveness dimensions, with robust predictive validity for attachment and child socioemotional outcomes. For further details on the coding procedure and coder training see Supplementary Materials.

##### Stress-Related Neural Activity

Given the limited sample size, we aimed to reduce the dimensionality of our MRI data through factor analysis. On our 10-cluster fMRI data (see Table 1), we first tested an a-priori two-factor confirmatory factor analysis (CFA) (Emotion vs Threat reactivity). Fit was poor (CFI=.71, RMSEA=.17), with several non-significant threat loadings (Full CFA statistics, eigenvalues, and loading matrices appear in the Supplementary Materials).

We therefore performed an exploratory principal component analysis (PCA) on the 10 clusters. Five orthogonal components explained 80 % of total variance. PC1 loaded strongly on emotion-condition clusters and was labelled Neural-Emotion. PC2 displayed modest to moderate cross-condition loadings (|λ|≈.30–.49), consistent with functional heterogeneity. It included the dorsal superior frontal gyrus (SFG) and midcingulate gyrus (MCG). Whereas the SFG is linked to top-down visuospatial attention and cognitive control (Eippert et al., 2007; Engen & Singer, 2015; Pallesen et al., 2009), the MCG is associated with attentional resource allocation under cognitive load and motivational conflict (Choi et al., 2014; Egner et al., 2005; Jueptner et al., 1997). We thus labelled PC2, Neural-Attentional Control. PC3 loaded on all and only threat-condition clusters and was thus labelled Neural-Threat. PC4 displayed moderate-to-strong cross-loadings (λ=.34–.67) on precuneus, associated with internally oriented mentation, rumination, and stress-related default mode network (DMN) modulation (Jacob et al., 2020; Soares et al., 2013) and on dorsal SFG. Given the plausible top-down influence of SFG on DMN states, we labeled PC4, Neural-Control/DMN Gating. PC5, dominated by a single loading on the same precuneus region (λ=.69), was labeled Neural-Preoccupation. All five components were entered as predictors in subsequent analyses.

**Table 2.** Loadings Matrix from Principal component (PC) analysis of 10 neural clusters.

| Cluster | Condition | PC1<br>(Emotion) | PC2<br>(Attentional<br>Control) | PC3<br>(Threat) | PC4<br>(Control/DMN) | PC5<br>(Preoccupation) |
| --- | --- | --- | --- | --- | --- | --- |
| Precentral Gyrus | Emotion | 0.451 |  |  |  |  |
| Inferior Frontal Gyrus | Emotion | 0.417 |  |  |  |  |
| Superior Frontal Gyrus | Emotion | 0.409 | 0.307 |  |  |  |
| Precentral Gyrus | Emotion | 0.451 |  |  |  |  |
| Precuneus | Emotion | 0.362 |  |  | 0.342 | 0.685 |
| Mid Cingulate | Threat |  | 0.497 | 0.350 |  |  |
| Superior Frontal Gyrus | Threat |  | 0.407 | 0.324 | 0.671 |  |
| Inferior Frontal Gyrus | Threat |  |  | 0.603 |  |  |
| Inferior Temporal Gyrus | Threat |  |  | 0.398 |  |  |
| Anterior Cingulate | Threat |  |  | 0.415 |  |  |
| **Explained Variance (%)** |  | **36.89** | **18.41** | **15.54** | **8.19** | **6.41** |

Loadings represent the contribution of each neural cluster to five components derived from PCA on z-scored beta values during the fMRI film task. These five orthogonal components explained ~ 85 % of total variance. PC1 (“Emotion”) indexes emotion-specific reactivity (i.e., prosocial/romantic + threatening > neutral); PC2 (“Attentional Control”) reflects fronto-cingulate control; PC3 (“Threat”) captures threat-specific reactivity (i.e., threatening > prosocial/romantic); PC4 (“Control/DMN”) aligns with control-default mode features; PC5 (“Preoccupation”) isolates precuneus activity. Cells list standardized loadings; blanks indicate |loading| < .30. The bottom row reports percent variance explained by each component. DMN = default mode network.

#### 2.4.2. Covariates

Given their established importance in the development of disruptive behaviors, we included household socioeconomic status (SES), child age, and child sex as covariates (Huisman et al., 2010; Theurel & Gentaz, 2018; Zahn-Waxler et al., 2008, respectively).

##### SES

Initial screening was conducted via the Geneva Sociodemographic Questionnaire (GSQ) (Sancho Rossignol et al., 2010), including a detailed overview of the parents’ SES, characteristics and history. SES was derived from the Swiss Largo Index; higher index values correspond to greater economic hardship (i.e., lower SES).

#### 2.4.3. Outcome variable

##### Disruptive Behaviors

To derive a composite outcome variable for child disruptive behaviors, we used the French version of the K-SADS-PL, which included attention-deficit/hyperactivity disorder (ADHD), conduct disorder (CD), oppositional defiant disorder (ODD), and disruptive mood dysregulation disorder (DMDD). This instrument has demonstrated strong reliability in previous research (Vandeleur et al., 2012). In the current study, we focused on selected modules of the interview, assessing the number of endorsed symptoms for each of the following disorders. The Disruptive Behavior composite was the sum of z-scored symptom counts from the ADHD, ODD, CD, and DMDD modules.

### 2.5. Data Analysis

#### 2.5.1. Attrition Analyses

Attrition analyses were conducted to determine if dropout (n=82) was related to demographic, psychosocial or clinical traits (see Attrition Analyses in the Methods of the Supplementary Materials for more details).

#### 2.5.2. Descriptive Statistics

Baseline characteristics were summarized separately for mothers without and with IPV-PTSS. Normality of each continuous variable was checked with the Kolmogorov-Smirnov test. Variables that met the assumption are reported as mean (M) ±□standard deviation (SD) and compared with independent-samples *t*-tests; variables that violated normality are reported as median□±□interquartile range (IQR) and compared with Mann-Whitney U-tests. Categorical variables are presented as *n* (%) and analyzed with chi-square (χ²) tests. All *p*-values are two-tailed. The composite score of Disruptive behavior was z-score transformed (M=0, SD=1).

#### 2.5.3. Bivariate associations (correlations)

To examine zero-order relations among study variables, we computed correlations between group status (non-IPV-PTSS vs. IPV-PTSS; coded 0/1), all predictors (P3 child age, SES, sex, PTSS severity, parental distress, maternal sensitivity, and the five neural components), and the outcome (Disruptive behaviors). Pearson’s *r* was used when both variables were approximately normal and continuous; otherwise Spearman’s ρ was used. Associations between the binary group indicator (0/1) and continuous variables were estimated with point-biserial correlations. Positive ρ indicates higher values in the group coded ‘1’. All tests were two-tailed (significance at α=.01 due to multiple comparisons).

#### 2.5.4. Model analysis strategy

In alignment with our first two empirical inquiries (i.e., whether maternal PTSS or neural reactivity relate to child disruptive behaviors), we modeled child disruptive behaviors using linear regression with forward stepwise selection under a Bayesian Information Criterion (BIC) stopping rule. Candidate predictors were PTSS Severity, Maternal Sensitivity, Parental Distress, Neural-Emotion, Neural-Attentional Control, Neural-Threat, Neural-Control/DMN, and Neural-Preoccupation. Socio-demographic covariates (SES, child age, child sex) were forced in all models.

Multicollinearity was checked considering VIFs < 5 as acceptable. Detailed diagnostics were performed on the fitted models.

#### 2.5.5. Moderation Analyses

In alignment with our third inquiry (i.e., whether maternal sensitivity moderates any neural-behavior link), we conducted moderation analyses, with Maternal Sensitivity as the moderating variable, on all significant neural components, independently, as per the maximally fitting model. Here, we again used forward stepwise regression with a BIC stopping rule. The interaction term was entered with both main effects of its two composite variables included in the model. Socio-demographic covariates (SES, child age, child sex) were forced in all models.

#### 2.5.6. Outliers Detection and Model Robustness

We assessed potentially influential outliers using QQ plot, Cook’ distance, leverage, and studentized residuals plot. Outlying observations were flagged as potentially influential if any screening rule was met: Cook’s distance > 4/*n*, leverage *h*_ii_ > 2*pred/*n*, or ∣studentized residual∣>3 (Fox, 1991), where *n* is the number of observations used in the model and *k* is the number of estimated coefficients (including the intercept). Because influential points can distort OLS estimates, we re-estimated the final linear models using robust M-estimation with Tukey’s bisquare (biweight) weights, implemented via iteratively reweighted least squares, which down-weights large residuals and yields estimates less sensitive to outliers and mild heteroscedasticity (cf. Holland & Welsch, 1977). Robust results are reported in the main text; conventional OLS fits and diagnostic plots appear in the Supplementary Materials.

#### 2.5.7. Statistical Power

Post-hoc power for the final (best-fitting) model was estimated from the observed *R^2^*(converted to Cohen’s *f^2^*) using the noncentral F-distribution at α=.05; full formulas and computation steps are provided in the Supplementary Materials.

## 3. Results

### 3.1. Attrition Analyses

Attrition analysis findings indicated that dropout was likely unrelated to measured demographic, psychosocial, or clinical variables, reducing the likelihood of systematic bias in the retained sample (see Attrition Analyses in Results of Supplementary Materials). However, in a non-significant logistic regression model (*p*=.122), Parental Distress reached significance (β=-0.07 *p*=.043), with higher parental distress associated with higher dropout.

### 3.2. Descriptive Statistics

Table 3 illustrates the sample characteristics, including socio-demographic, psychological, behavioral and neurobiological traits. Relative to non-IPV-PTSS, the IPV-PTSS group reported significantly increased PTSS (*p*<.001) and Parental Distress (*p*<.001) as well as lowered Maternal Sensitivity (*p*=.001). Additionally, the IPV-PTSS group showed higher Neural-Control/DMN scores (*p*=.022), indicating stronger co-recruitment of dorsal SFG (control) with precuneus (preoccupation/DMN) during stress-related processing, consistent with greater top-down modulation of internally oriented processing.

**Table 3.** Sample characteristics.

| Variable | Total<br>(Mean ± SD) | Non-IPV-PTSS<br>(Mean ± SD) | IPV-PTSS<br>(Mean ± SD) | p-value |
| --- | --- | --- | --- | --- |
| N | 28 | 14 (50%) | 14 (50%) | 0.590 |
| P3 Child Age | 10.86 ± 1.56 | 10.71 ± 1.44 | 11.00 ± 1.71 | 0.865 |
| SES | 5.25 ± 1.99 | 4.64 ± 1.98 | 5.86 ± 1.88 | 0.121 |
| Sex | 11F, 17M | 6F, 8M | 5F, 9M | 0.925 |
| PTSS Severity | 52.54 ± 38.40 | 19.00 ± 6.42 | 86.07 ± 24.47 | <.001 |
| Parental Distress | 15.25 ± 8.81 | 9.00 ± 4.72 | 21.50 ± 7.40 | <.001 |
| Maternal Sensitivity | 5.25 ± 1.38 | 6.07 ± 1.07 | 4.43 ± 1.16 | 0.001 |
| Neural Emotion | -0.15 ± 1.90 | -0.62 ± 1.89 | 0.31 ± 1.87 | 0.105 |
| Neural Threat | 0.22 ± 0.90 | 0.11 ± 0.66 | 0.32 ± 1.11 | 0.864 |
| Neural Attentional Control | -0.00 ± 1.41 | 0.05 ± 1.33 | -0.06 ± 1.55 | 0.671 |
| Neural Control/DMN Gating | -0.03 ± 0.93 | -0.40 ± 0.82 | 0.34 ± 0.91 | 0.022 |
| Neural Preoccupation | 0.02 ± 0.75 | 0.03 ± 0.84 | 0.02 ± 0.67 | 0.848 |
| Disruptive Behaviors | -0.40 ± 2.67 | -0.95 ± 3.17 | 0.15 ± 2.04 | 0.196 |

Note. All group differences were assessed with independent-samples t-tests (normality-tested, variance-adjusted, FDR-corrected). Sample size and sex differences between groups were analyzed using chi-square. IPV=interpersonal violence, F=females, M=males, P3=phase 3, SD=standard deviation, SES=socio-economic status ; DMN=default mode network.

### 3.3. Correlations

Table 4 illustrates bivariate correlations between Group (non-IPV/IPV), aforementioned predictors, covariates and the outcome variable (disruptive behaviors). Notably PTSS correlated positively with Parental Distress (*r*=.65, *p*<.001). Maternal Sensitivity correlated negatively with PTSS (*r*=-0.55, *p*<.001), Parental Distress (*r*=-.52, *p*<.01) and Neural-Emotion (*r*=-0.48, *p*=.010).

**Table 4.** Correlations.

|  | 1 | 2 | 3 | 4 | 5 | 6 | 7 | 8 | 9 | 10 | 11 | 12 | 13 |
| --- | --- | --- | --- | --- | --- | --- | --- | --- | --- | --- | --- | --- | --- |
| 1 Group | -- |  |  |  |  |  |  |  |  |  |  |  |  |
| 2 Phase 3 Child Age | 0.07 | -- |  |  |  |  |  |  |  |  |  |  |  |
| 3 SES | 0.17 | -0.21 | -- |  |  |  |  |  |  |  |  |  |  |
| 4 Sex | -0.06 | 0.25 | 0.20 | -- |  |  |  |  |  |  |  |  |  |
| 5 PTSS Severity | 0.81*** | 0.00 | 0.24 | -0.12 | -- |  |  |  |  |  |  |  |  |
| 6 Parental Distress | 0.71*** | 0.03 | 0.11 | -0.05 | 0.65*** | -- |  |  |  |  |  |  |  |
| 7 Maternal Sensitivity | -0.57*** | -0.30 | -0.12 | 0.24 | -0.55*** | -0.52** | -- |  |  |  |  |  |  |
| 8 Neural Emotion | 0.29 | 0.19 | 0.23 | 0.10 | 0.27 | 0.38 | -0.48** | -- |  |  |  |  |  |
| 9 Neural Threat | -0.08 | 0.12 | -0.01 | 0.02 | 0.05 | -0.04 | -0.12 | 0.00 | -- |  |  |  |  |
| 10 Neural Attentional Control | -0.04 | 0.22 | -0.04 | 0.19 | -0.13 | -0.01 | 0.34 | 0.00 | 0.00 | -- |  |  |  |
| 11 Neural Control/DMN | 0.38 | -0.11 | 0.02 | 0.39 | 0.19 | 0.23 | -0.16 | 0.00 | 0.00 | 0.00 | -- |  |  |
| 12 Neural Preoccupation | -0.01 | 0.21 | -0.10 | 0.07 | -0.08 | -0.08 | -0.11 | 0.00 | 0.00 | 0.00 | 0.00 | -- |  |
| 13 Disruptive Behaviors | 0.20 | -0.04 | -0.03 | 0.18 | 0.02 | -0.03 | -0.30 | 0.03 | -0.11 | -0.07 | 0.29 | 0.20 | -- |

Note. PTSS= post-traumatic stress symptoms; SES=socio-economic status measured via the Swiss Largo Index, where increasing SES reflects increasing economic hardship (i.e., lower SES). \*\*p≤.01, \*\*\*p<.001

### 3.4. Model Analyses

The robust M-estimation of the winning BIC-selected model was highly significant (*n*=28; *F*(7,20)=6.83, *R*²=.705; adj. *R*²=.602, *p*<.001, power > .99). Lower Disruptive Behaviors was associated with higher Maternal Sensitivity (β=-1.29, *p*<.001), higher Neural-Threat (β=-1.13, *p*=.007), older age (β=-0.90, *p*=.003), male sex (β=3.21, *p*<.001) and lower Neural-Preoccupation (β=1.37, *p*=.008) (see Table 5).

**Table 5.**
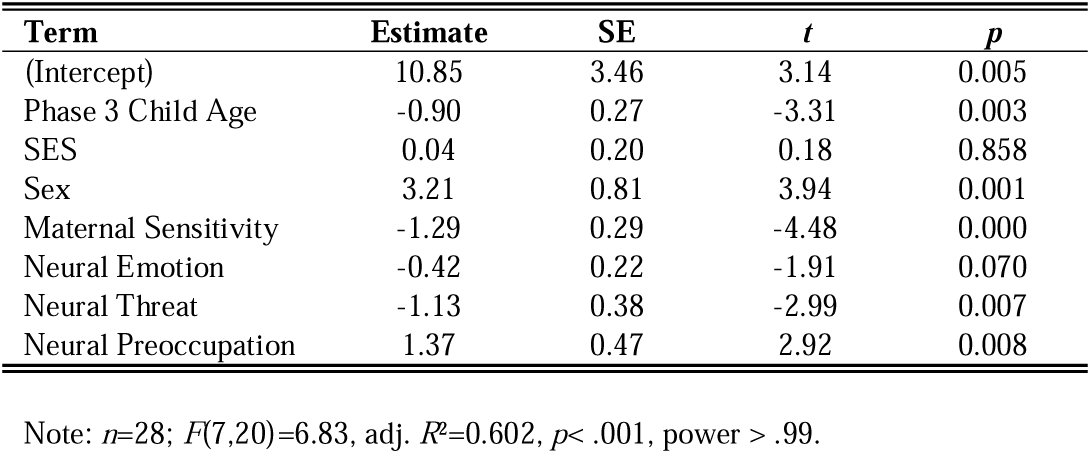
Robust M-Estimation of winning BIC-selected model for Disruptive Behaviors.

| Term | Estimate | SE | *t* | *p* |
| --- | --- | --- | --- | --- |
| (Intercept) | 10.85 | 3.46 | 3.14 | 0.005 |
| Phase 3 Child Age | -0.90 | 0.27 | -3.31 | 0.003 |
| SES | 0.04 | 0.20 | 0.18 | 0.858 |
| Sex | 3.21 | 0.81 | 3.94 | 0.001 |
| Maternal Sensitivity | -1.29 | 0.29 | -4.48 | 0.000 |
| Neural Emotion | -0.42 | 0.22 | -1.91 | 0.070 |
| Neural Threat | -1.13 | 0.38 | -2.99 | 0.007 |
| Neural Preoccupation | 1.37 | 0.47 | 2.92 | 0.008 |

Note: $n=28$ ; $F(7,20)=6.83$ , adj. $R^2=0.602$ , $p < .001$ , power $> .99$ .

### 3.5. Moderation Analyses

The robust M-estimation of the winning BIC-selected interaction model was significant (*n*=28; *F*(8,19)=10.91, *R*²=.746, *p*<.001, power > .99) and contained the Maternal Sensitivity × Neural-Threat term, which was significantly associated with Disruptive Behaviors (β=−0.95, *p*=.005). Main effects were consistent with the main-effects model: Age (β=−1.07, *p*=.002), Sex (β=3.56, p=.002), Neural-Threat (β=-1.72, *p*<.001), Neural Preoccupation (β=1.696, *p*=.001); Maternal Sensitivity and Neural-Emotion were marginally significant ((β=-0.72, *p*=.074 and β=-0.32, *p*=.080, respectively) (see Table 6).

**Table 6.** Robust M-Estimation of winning BIC-selected interaction model for Disruptive Behaviors.

| Term | Estimate | SE | *t* | *p* |
| --- | --- | --- | --- | --- |
| (Intercept) | 7.28 | 3.01 | 2.42 | 0.026 |
| Phase 3 Child Age | -0.92 | 0.21 | -4.31 | 0.000 |
| SES | 0.25 | 0.17 | 1.49 | 0.152 |
| Sex | 2.64 | 0.66 | 3.99 | 0.001 |
| Maternal Sensitivity | -0.61 | 0.32 | -1.91 | 0.071 |
| Neural Emotion | -0.32 | 0.18 | -1.85 | 0.080 |
| Neural Threat | 3.17 | 1.38 | 2.30 | 0.033 |
| Neural Preoccupation | 1.54 | 0.38 | 4.08 | 0.001 |
| Maternal Sensitivity × Neural Threat | -0.95 | 0.30 | -3.21 | 0.005 |

Maternal Sensitivity as a Moderator of Neural-Threat Reactivity and Child Disruptive behaviors, using Robust M-Estimation. Note: $n=28$ ; $F(8,19)=10.91$ , adj. $R^2=0.746$ , $p < .001$ , power $> .99$ .

#### 3.5.1. Post-hoc simple slopes analysis

To further examine the direction of the moderation effect, we conducted a simple slopes analysis at ±1 SD of Maternal Sensitivity, using a continuous moderation framework appropriate for our modest sample size and using robust M-estimation modeling. As shown in Figure 1, the association between maternal Neural-Threat activity and Disruptive Behaviors was significantly negative at higher levels of maternal sensitivity (β=-1.91, *t*(24) = −2.01, *p*=.044) (Table S5), but non-significant at lower levels (p=.701). A robust Johnson–Neyman analysis indicated that the simple slope of Neural Threat predicting Disruptive Behaviors was significant for moderate levels of Maternal Sensitivity (−0.03 to 1.35 SDs relative to the sample mean), but not at very low or very high levels of Maternal Sensitivity (Figure S1). Within this region the slope was negative, indicating that higher Neural-Threat was associated with lower Disruptive Behaviors when Maternal Sensitivity was in this range. These results suggest that elevated maternal threat-related neural activity likely associates with reduced child disruptive behaviors only in the context of moderate to high maternal sensitivity.

**Figure 2.**
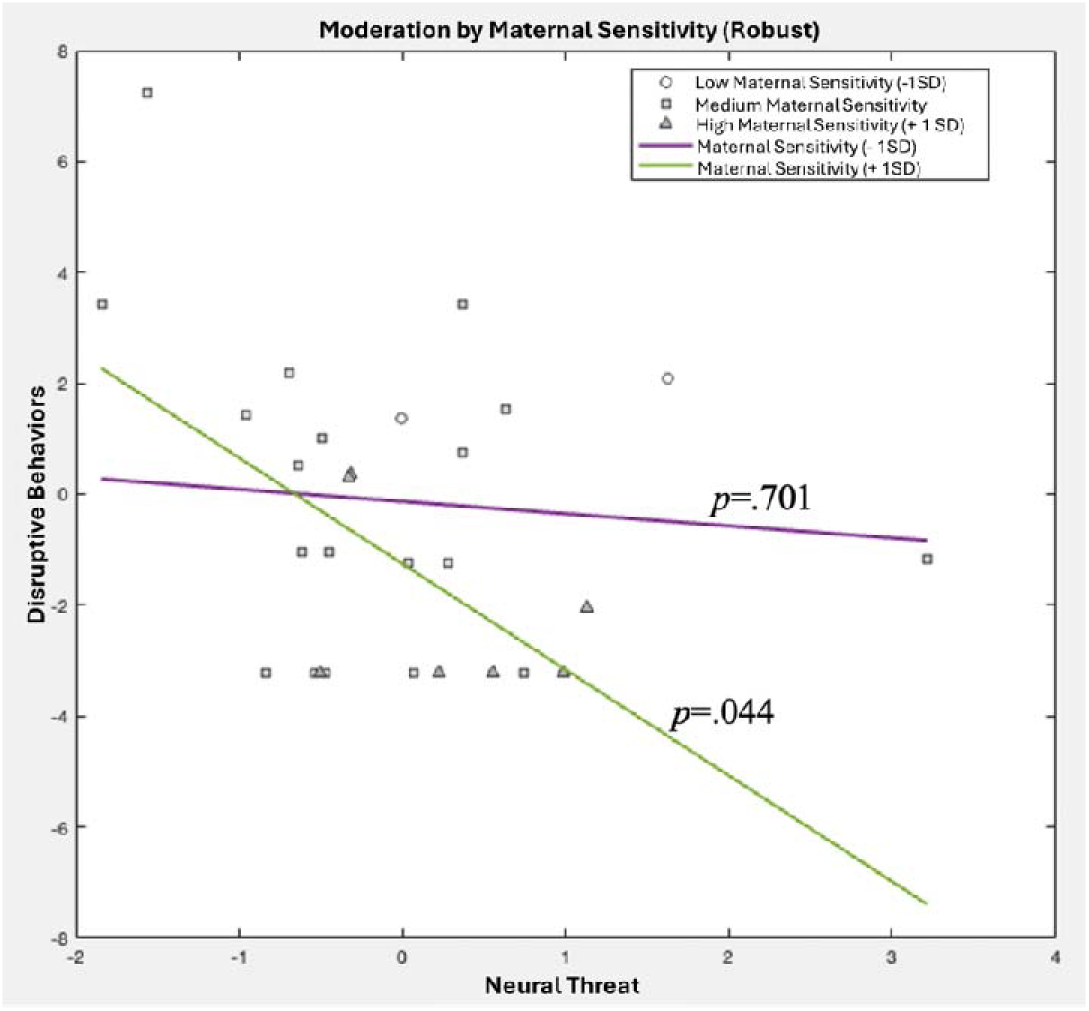
Moderation of Neural Threat by Maternal Sensitivity, using robust M-Estimation model.

## 4. Discussion

Youth violence is an urgent public-health problem: worldwide, an estimated 193,000 homicides occur each year among people aged 15–29 (World Health Organization, 2024), and public health data indicate rising aggression and weapon-related injuries (Gwata et al., 2024), making adolescence a critical window for prevention. Because aggressive trajectories often strengthen in the peri-pubertal years, we focused on disruptive behaviors, that is, persistent impulsivity, rule-breaking, and defiance.

Disruptive-behavior pathways are central to intergenerational trauma, forecasting later violence, criminality, and revictimization, and they are overrepresented among children of parents exposed to IPV. Given the high global prevalence of IPV, particularly among women, understanding how maternal IPV-related trauma shapes offspring behavior is a public-health priority. Guided by this rationale, our primary aim was exploratory and data-driven: to identify which maternal factors best account for variance in peri-pubertal disruptive behaviors. We examined whether maternal IPV-related PTSS severity relates to child disruptive behavior, whether maternal stress-related neural reactivity in toddlerhood contributes unique variance, and whether maternal sensitivity moderates any neural–behavior associations, with higher sensitivity expected to buffer risk. These analyses were hypothesis-generating and intended to provide testable predictions for future, adequately powered confirmatory studies.

Contrary to some reviews of the literature (e.g., Van Ee et al., 2016), maternal PTSS severity did not show a direct association with child disruptive behaviors in our multivariable models. This pattern aligns with growing evidence that PTSS-related risk is often indirect and embedded in contextual stress and caregiving processes. In a large, well-powered study of asylum-seeking dyads, Bachem and colleagues (2024) reported that post-migration living difficulties and complex-PTSD features (e.g., disturbances in self-organization), rather than PTSS alone, accounted for intergenerational transmission leading to disruptive behaviors. Other reports have found PTSS–child behavior links that may reflect correlated mechanisms or third variables, such as HPA-axis–related methylation (NR3C1), albeit in small samples (Cordero et al., 2022), or pathways involving paternal violence with maternal PTSS as a mediator (Schechter & Rusconi, 2011).

Parenting behavior also appears likely to confound PTSS–child behavior associations (Brady et al., 2024; Greene et al., 2018; Thompson et al., 2024). Our models simultaneously adjusted for maternal sensitivity and socio-demographics and included neural indices. Under this specification, PTSS contributed no unique variance. Thus, our null main effect is consistent with integrative models in which PTSS likely elevates risk primarily through contextual stressors and parenting behavior, not as an isolated predictor. Even so, our sample is modest; small direct effects cannot be ruled out and should be tested in larger, multi-method cohorts that also consider biomarkers and paternal factors (e.g., Brady et al., 2024; Suardi et al., 2020).

Although we observed no direct effect of PTSS, maternal neural stress-related reactivity (i.e., Neural-Threat, Neural-Preoccupation) illustrated a direct association with disruptive behaviors in the peri-pubertal period, when adjusting for maternal sensitivity. This supports the notion that while maternal neurobiological stress responses to threat- and/or emotion-specific information may be directly associated with child behavioral regulation, maternal caregiving may play a moderating role. Indeed, moderation analyses demonstrated that maternal sensitivity did significantly modulate the relationship between neural threat-specific activity and child disruptive behaviors; higher maternal neural threat-specific reactivity was associated with fewer disruptive behaviors, but only when maternal sensitivity was also high. While preliminary, these results underscore the importance of both neural and relational processes in shaping long-term child outcomes.

### 4.1. Threat-related neural activity

To examine the role of maternal stress-related brain function in intergenerational risk, this study applied Principal Component Analysis (PCA) to maternal neural responses across ten regions previously linked to emotion and threat processing. This allowed for a systems-level view of stress-related reactivity, capturing broader neural dynamics beyond isolated regions. Whereas five principal components were extracted, two key components emerged: Neural-Threat (corresponding to threat-specific neural activation patterns) and Neural-Preoccupation (corresponding to emotion-specific precuneus activity).

Maternal neural reactivity during toddlerhood, captured by both Neural-Threat and Neural-Preoccupation, was associated with disruptive behaviors in children at peri-puberty. That is, higher threat-specific neural activity predicted fewer disruptive behaviors, suggesting a potential protective role. In contrast, elevated emotion-specific and preoccupation-related precuneus activity was linked to increased disruptive behaviors, indicating a potential risk factor.

The threat-specific neural reactivity component, Neural-Threat, included five cortical regions: MCG, SFG, inferior frontal gyrus (IFG), inferior temporal gyrus (ITG), and pregenual anterior cingulate cortex (ACC), regions implicated in top-down regulation and attentional control in response to threat (Choi et al., 2014; Clarke & Johnstone, 2013; Egner et al., 2005; Han et al., 2008; Jueptner et al., 1997). For instance, the MCG and SFG coordinate attentional resources under cognitive load or motivational conflict (Choi et al., 2014; Egner et al., 2005; Jueptner et al., 1997) while the IFG facilitates affective inhibition (Clarke & Johnstone, 2013; Han et al., 2008) and cognitive flexibility (Friedman & Robbins, 2022). The ITG and ACC contribute to anticipatory threat detection (Cornwell et al., 2007; Mohanty et al., 2009) and integration of emotional salience and interoception (Murray et al., 2012; Palomero-Gallagher et al., 2019).

The association between Neural-Threat and disruptive behaviors was further moderated by maternal sensitivity: specifically, high maternal sensitivity amplified the inverse relationship between threat-specific reactivity and child disruptive behaviors. No moderation effect was found for Neural-Preoccupation in the best fitting model. These preliminary findings suggest that stress-responsive maternal brain function may interact with caregiving behaviors to shape child developmental trajectories.

Interestingly, prior findings (Moser et al., 2015) linked increased activity in MCG, SFG, and IFG with higher maternal sensitivity, whereas ITG and ACC were negatively associated with sensitivity. This balance suggests that Neural-Threat captures both adaptive and dysregulated responses to threat. When paired with high maternal sensitivity, this neural profile may enhance emotion regulation and attentional control during stressful interactions, thereby reducing child behavioral dysregulation. This highlights the potential regulatory role of sensitive caregiving in mitigating behavioral risk.

### 4.2. Neural-Preoccupation (Precuneus)

Beyond threat-specific neural reactivity, we observed that increased activity in maternal emotion-specific and preoccupation-related precuneus during toddlerhood significantly predicted greater disruptive behaviors in children at peri-puberty. The precuneus is considered a hub of the DMN (Utevsky et al., 2014) involved in internally directed thought, self-referential processing, and memory-based simulation (Cavanna, 2007).

Prior research has implicated precuneus hyperactivity in cognitively demanding tasks involving mental imagery (e.g., mental scanning from text [Mellet, 2002]) and risky decision-making under cognitive load (Gathmann et al., 2014), functions relevant to stress sensitivity. Not surprisingly, the precuneus is associated with rumination (i.e., repetitive thinking patterns and focus on negative states [Jacob et al., 2020]). Notably, precuneus hyperactivation has also been reported in PTSD patients during flashback/reliving states compared to dissociative states (Lanius et al., 2005), and in resting-state fMRI studies examining default mode alterations under acute stress (Soares et al., 2013), suggesting its involvement in trauma-related cognitive-emotional processing.

In the context of this literature, maternal hyperactivation in this region may signal heightened internally focused attention, stress-related rumination, or impaired disengagement from emotionally salient cues during early caregiving. These traits may reduce maternal attunement or increase stress contagion within the parent-child dyad, thereby elevating the child’s risk for behavioral dysregulation.

Although based on a single region, the precuneus may thus represent a meaningful neurobiological marker of intergenerational vulnerability, warranting further investigation in larger samples and more refined network-based analyses.

### 4.3. Clinical implications

These findings underscore the importance of addressing both maternal IPV-related psychopathology, the quality of the mother–child relationship, and the relation between the two factors during early sensitive periods of development. Intervening during the first four years of life, when the psychobiological systems that contribute together to the development of children’s self-regulation of emotion, arousal, and aggression, are working intensively and rapidly together, and during which time, parents are often more receptive to change, may help limit the emergence of disruptive behavior in children. Enhancing maternal sensitivity during this formative period not only supports mutual regulation as a prerequisite to self-regulation but may complement adaptive child response to threat at neural and psychological levels.

Notably, our results suggest that maternal threat-specific neural activity, when paired with high sensitivity, may buffer against child disruptive behaviors. This implies that strengthening mothers’ ability to detect and regulate responses to perceived interpersonal threat, particularly when triggered by child distress or defiance, may improve caregiving behavior, reduce reactivity, and possibly, in the long-run, intergenerational transmission of violence when their children become adults and parents. Trauma-informed interventions that refine maternal interpretations of child behavior and strengthen top-down regulation, integrating psychodynamic, imaginal exposure, body-oriented, attachment- and mentalization-based approaches with video feedback (Schechter, 2024), and that explicitly target stress-related rumination (e.g., rumination-focused cognitive-behavioral therapy, metacognitive strategies, mindfulness-based cognitive therapy, imagery rescripting) may enhance maternal sensitivity, reduce ruminative preoccupation, and improve child outcomes. These findings support a more integrative, neurobehavioral model of parenting intervention in trauma-exposed families.

### 4.4. Limitations

The neural clusters used in this analysis were originally derived based on their associations with maternal sensitivity (Moser et al., 2015). However, the current study avoids circularity by testing a distinct outcome, child psychopathology at peripuberty, and by reducing neural data through PCA to extract broader patterns of reactivity. This is further supported by the significant association of Neural-Threat and Neural-Preoccupation components with disruptive outcomes, even when accounting for maternal sensitivity. Furthermore, the mixed valence of original associations (both positive and negative) within threat-specific clusters (Moser et al., 2015) reduces the risk of directional bias in the composite Neural-Threat score. Although PCA helped identify coherent, functionally interpretable neural patterns, moderate cross-loadings in some clusters limited component specificity, a known limitation of the method (Costello & Osborne, 2005). These cross-loadings nonetheless attest to the functional heterogeneity that is evidenced in individual brain regions, such as the inferior frontal gyrus (Clos et al., 2013).

Attrition did not appear random, i.e., families with higher parental distress were more likely to drop out, indicating informative missingness and potential selection bias. Consequently, our findings should be interpreted cautiously and not overgeneralized beyond the retained sample, especially given the modest sample size.

Finally, despite the elevated power measured in our winning stepwise models illustrating main effects of neural stress-related reactivity, this study was limited by its reliance on principal components rather than whole-brain or region-specific imaging analyses. Replication in larger, independent cohorts, using these broader imaging analysis methods is warranted to confirm the robustness of this effect.

### 4.5. Conclusion

The present findings suggest that maternal threat-specific neural reactivity, especially when coupled with higher maternal sensitivity, supports children’s behavioral regulation, whereas precuneus-dominant activity may signal risk. Together, they underscore the need to consider psychological and neurobiological dimensions of maternal functioning and their convergence in caregiving. While preliminary and based on a modest, selective sample, the results point to early, trauma-informed interventions that reduce maternal distress, promote sensitive caregiving, and support adaptive stress-response processes to foster healthier child development.

## Data Availability

The data that support the findings of this study are available from the corresponding author, RJM, upon reasonable request

## 7. Declaration of generative AI and AI-assisted technologies in the manuscript preparation process

During manuscript preparation, the authors used ChatGPT (OpenAI; GPT-4/5) to improve wording and to assist with drafting/optimizing statistical code. The authors reviewed, edited, and verified all AI-assisted text and code, conducted all analyses themselves, and take full responsibility for the content. No identifiable data were entered into the tool.

## Acknowledgments

This research reported in this paper was made possible by funding from Swiss National Science Foundation grants [numbers 32513B_204863 and 51AU40_125759] as well from la Fondation Prim’Enfance, Geneva, to Professor Daniel Schechter. We thank Ms. Sondes Jouabli for conducting the clinical interviews and contributing to data collection. We also acknowledge Ms. Barbara Garrido Araujo and Ms. Monica Lourido Garcia in the preparation of this manuscript. We additionally thank Dr. Dominik A. Moser for his help in the initial conceptual planning of this paper. This work would not have been possible without the families, mothers and children who maintained their participation in the study over a period of twelve years.

