## Supplemental Materials for "Maternal Stress-Related Neural Reactivity and Caregiving Sensitivity in Early Childhood: Associations with Peri-Pubertal Disruptive Behaviors"

### 1. Supplementary Materials

#### 1.1. Methods

##### 1.1.1. **Measure**

###### 1.1.1.1. **Maternal Sensitivity (CARE Index)**

Maternal sensitivity was measured using the CARE-Index (Crittenden & Bonvillian, 1984; Kunster et al., 2010), a widely used observational tool that assesses parental sensitivity, control, and unresponsiveness during short mother–infant/toddler interactions. The interaction task involved 5 minutes of unstructured free-play between mother and child (aged 12–44 months) using a standardized toy set (e.g., blocks, dolls). Mothers were instructed to "play as you normally would." Videos were recorded just before the child’s MRI session.

Two doctoral-level psychologists, blind to clinical group status, coded the videos using the CARE-Index’s 8-item coding scheme, which includes facial, vocal, bodily, and sequential indicators. Coders were reliability-trained by completing the standard CARE-Index certification (approximately 40 hours of instruction and 20 reliability videos). Twenty-five percent of sessions were double-coded; inter-rater reliability for the sensitivity scale was excellent (ICC [2,1]=0.86). Discrepancies exceeding 1 point were resolved by best estimate consensus.

CARE-Index raw sensitivity scores (0–14), with 0 being extremely insensitive, 7 being normally sensitive, and 14 being exceptionally sensitive were used continuously and are strongly predictive of child attachment security and socioemotional development (Crittenden, 2006). In our sample, internal consistency for sensitivity was good (Cronbach’s α=.82).

##### 1.1.2. Normality & Transformation

Each variable’s distribution was examined with Kolmogorov–Smirnov tests. Variables that violated normality were converted to tied ranks and then z-standardized to preserve metric comparability. All child-symptom items were subsequently z-standardized and averaged to form the Disruptive behavior composite.

##### 1.1.3. Group Comparisons

Independent-samples *t*-tests compared IPV-exposed and control mothers on age, SES, PTSS, MDDS, parenting stress, neural components, and child outcomes. Levene’s tests confirmed equal variances; when violated, Welch’s correction was applied. Multiple comparisons were controlled via the Benjamini–Hochberg false-discovery-rate (FDR) procedure (*q* < .10).

##### 1.1.4. Correlation Matrix & Collinearity

Bivariate associations were displayed in a full correlation matrix: Pearson’s *r* for normally distributed pairs, Spearman’s ρ when either variable remained non-parametric post-transformation. Variance Inflation Factors (VIFs) were computed for each predictor by regressing it on all others; all fell below 5 (range=1.21–4.34), indicating no multicollinearity concerns (Table S1).

##### 1.1.5. Confirmatory Factor Analysis Full Results

To explore the underlying structure of neural cluster data and reduce dimensionality, we first tested an a-priori two-factor measurement model R via lavaan (v0.6-19) using maximum likelihood estimation. Ten neural cluster volumes (C1–C10) were specified to load on two correlated latent factors: Emotion-Specific Reactivity (C1–C5) and Threat-Specific Reactivity (C6–C10). Factor variances were fixed to 1 for identification. Model fit was evaluated via χ², CFI, TLI, RMSEA (with 90% CI), and SRMR.

Our confirmatory two-factor model failed to achieve acceptable fit (CFI<.90, RMSEA>.10), and several loadings on the Threat factor were non-significant, justifying the use of exploratory PCA to derive data‐driven components (see Supplementary Materials for full CFA results).

The two-factor CFA exhibited significant misfit: χ²(34)=49.84, p=.039; CFI=.858; TLI=.812; RMSEA=.123 (90% CI [.029, .192], p (RMSEA≤.05)=.078, p (RMSEA≥.08)=.829); SRMR=.137. Although C1–C5 loaded strongly on the Emotion-specific factor (standardized λ=.80–.88, p<.001), several Threat-Specific factor loadings were weak or non-significant (e.g., C7: λ=.40, p=.056; C8: λ=.37, p=.070; C9–C10 non-significant) and C6’s residual variance was negative. The inter-factor correlation was near zero (r=−.10, p=.567), and multiple modification indices suggested cross-loadings. These indicators confirm that the hypothesized structure poorly captures the observed covariance.

##### 1.1.6. Principal component Analysis

Given the null findings for our CFA we then conducted an exploratory Principal Component Analysis (PCA) on data from 10 neural regions of interest (ROIs). The ROIs corresponded to clusters extracted from neuroimaging data related to emotional and neutral conditions (Table 1). Before performing PCA, all neural data were standardized using z-scores to ensure that each variable contributed equally to the analysis.

The PCA was executed to transform the original set of variables (ROIs) into a smaller set of uncorrelated principal components that captured the most variance in the dataset. Eigenvalues were extracted to assess the amount of variance explained by each principal component, and cumulative explained variance was calculated to identify how many components should be retained. A common threshold of 80% cumulative variance was used to determine the number of principal components for further analysis.

##### 1.1.7. Attrition Analyses

To evaluate whether differential dropout biased longitudinal findings, we compared families who completed both Phase 1 (P1; N=113) and Phase 3 (P3; n=28) with those lost to follow-up (n=85; 75.22 %). Predictors included P1 child age, maternal age, maternal violent trauma history, P1 Parental Distress, and P1 child behavioral dysregulation.

Maternal violent trauma, i.e., physical or sexual abuse exposure from childhood onward, was measured via a semi-structured interviewed using the Brief Physical and Sexual Abuse Questionnaire (BPSAQ, (Marshall et al., 1998)). Parental Distress at P1 was assessed using the Parental Distress subscale of the Parenting Stress Index–Short Form (PSI-SF) (Abidin, 1995). Child behavioral dysregulation was assessed via the maternal-reported Infant-Toddler Social Emotional Assessment (ITSEA, (Carter et al., 2003)).

All continuous variables were initially screened for normality (Kolmogorov–Smirnov). Parametric variables were examined with independent-samples *t*-tests (equal-variance assumption checked); non-normal variables (socio-economic status, maternal abuse history) were compared with Mann-Whitney *U* tests. Sex differences were assessed with χ². Finally, a multivariate logistic regression entered all baseline variables simultaneously to test their joint ability to predict study completion.

#### 1.2. Results

##### 1.2.1. Attrition Analyses

Completers and dropouts (scored as 1 and 0, respectively) did not differ on any baseline characteristic. Child age, maternal age, dysregulation, and parental distress all showed nonsignificant *t* values (|*t*| ≤ 1.07, *ps*>.20). Median SES and maternal physical/sexual abuse scores were identical across groups (Mann-Whitney *z*=0, *p*=1.00). The proportion of boys and girls was equivalent (χ²(1)=0.02, *p*=.88). Logistic regression confirmed that the combined set of predictors did not distinguish completers from dropouts (χ²(7)=11.40, *p*=.122); Parental Distress reached significance (*β*=-0.07 *p*=.043), with higher Parental Distress associated with higher attrition. Because the overall model was not significant, and direct group differences in Parental Distress yielded non-significant results, this association should be interpreted cautiously. These findings indicate that attrition was likely unrelated to measured demographic, psychosocial, or clinical variables, reducing the likelihood of systematic bias in the retained sample.

##### 1.2.2. Original Models

###### 1.2.2.1. Main Effects

The original OLS BIC-selected model (*n*=28, *F*(7,20)=7.18, *R*²=.715; adj. *R*²=.616, *p<*.001) was highly significant. Higher Maternal Sensitivity (*β*=-1.74, *p*<.001) and higher Neural-Threat (*β*=-1.38, *p*=.007) were associated with lower Disruptive scores; older age (*β*=-1.36, *p*=.003) with lower Disruptive; male sex (*β*=3.23, *p*<.001) and higher Preoccupation (*β*=1.14, *p*=.005) with higher Disruptive behavior. Neural-Emotion was marginally significant (*β*=-0.78, *p*=.068).

| **Term** | **Estimate** | **SE** | ***t*** | ***p*** |
| --- | --- | --- | --- | --- |
| (Intercept) | -2.09 | 0.57 | -3.67 | 0.002 |
| P3 Child Age | -1.36 | 0.41 | -3.34 | 0.003 |
| SES | 0.07 | 0.37 | 0.18 | 0.858 |
| Sex | 3.23 | 0.79 | 4.09 | 0.001 |
| Maternal Sensitivity | -1.74 | 0.39 | -4.52 | 0.000 |
| Neural Emotion | -0.78 | 0.41 | -1.93 | 0.068 |
| Neural Threat | -1.38 | 0.46 | -3.03 | 0.007 |
| Preoccupation | 1.14 | 0.36 | 3.13 | 0.005 |
| **Table S2.** Maximally fitting OLS model using forward stepwise regression and BIC as a stopping rule. Note: *n*=28; *F*(7,20)=7.18, adj. *R*²=0.616, *p*<.001, power > .99 | | | | |

###### 1.2.2.2. Moderation Analysis

The winning BIC-selected interaction model (*n*=28, *F*(8,19)=9.75, *R*²=.804; adj. *R*²=.722, *p<*.001) was highly significant and contained the Maternal Sensitivity × Neural-Threat term, which was also significant (*β*=-0.91, *p*=.008). Main effects were consistent with the main-effects model: Age (*β*=-0.89, *p*<.001), Sex (β=2.77, p<.001), Neural-Threat (*β*=-1.74, *p*<.001), Preoccupation (*β*=1.65 *p*<.001); Neural-Emotion and Maternal Sensitivity were not significant (*p*s>.05).

| **Term** | **Estimate** | **SE** | ***t*** | ***p*** |
| --- | --- | --- | --- | --- |
| (Intercept) | 6.67 | 3.14 | 2.12 | 0.047 |
| Phase 3 Child Age | -0.89 | 0.22 | -3.98 | 0.001 |
| SES | 0.25 | 0.18 | 1.40 | 0.179 |
| Sex | 2.77 | 0.69 | 4.01 | 0.001 |
| Maternal Sensitivity | -0.58 | 0.33 | -1.72 | 0.101 |
| Neural Emotion | -0.31 | 0.18 | -1.71 | 0.103 |
| Neural Threat | 3.02 | 1.44 | 2.09 | 0.050 |
| Preoccupation | 1.65 | 0.40 | 4.18 | 0.001 |
| Maternal Sensitivity × Neural Threat | -0.91 | 0.31 | -2.94 | 0.008 |
| **Table S3.** OLS-based maximally fitting moderation effects model using forward stepwise regression and BIC as a stopping rule. Note: *n*=28; *F*(8,19)=9.75, adj. *R*²=0.722, *p*<.001, power > .99 | | | | |

##### 1.2.3. Simple Slopes Metrics

| Slope | | |  | Standard Error | | |  | *t* | | |  | *p* | | |
| --- | --- | --- | --- | --- | --- | --- | --- | --- | --- | --- | --- | --- | --- | --- |
| M=+ 1SD |  | M=– 1SD |  | M=+ 1SD |  | M=– 1SD |  | M=+ 1SD |  | M=– 1SD |  | M=+ 1SD |  | M=– 1SD |
| -1.910 |  | -0.219 |  | 0.949 |  | 0.571 |  | -2.013 |  | -0.384 |  | 0.044 |  | 0.701 |
| Table S5. Simple Slopes (±1 SD). X=Neural Threat, M=Maternal Sensitivity. | | | | | | | | | | | | | | |

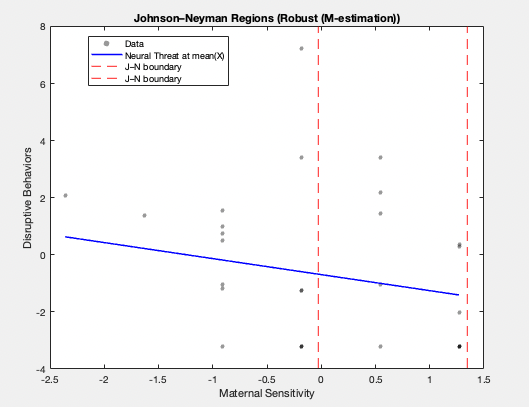

Figure S1. Johnson-Neyman plot examining the association between Neural-Threat and Disruptive behaviors, as moderated by Maternal Sensitivity. Neural-Threat and Maternal Sensitivity are mean-centered.

##### 1.2.4. Diagnostic Plots

###### 1.2.4.1. Main Effects

Regression diagnostics from the original OLS model flagged two observations, one case with |studentized residuals| > 3 (i.e., 3.18) (Figure S2A) and one case with elevated Cook’s D (0.257, with threshold at 0.143) (Figure S2C) and elevated leverage (0.592 with threshold at 0.571) (see Figure S2D).

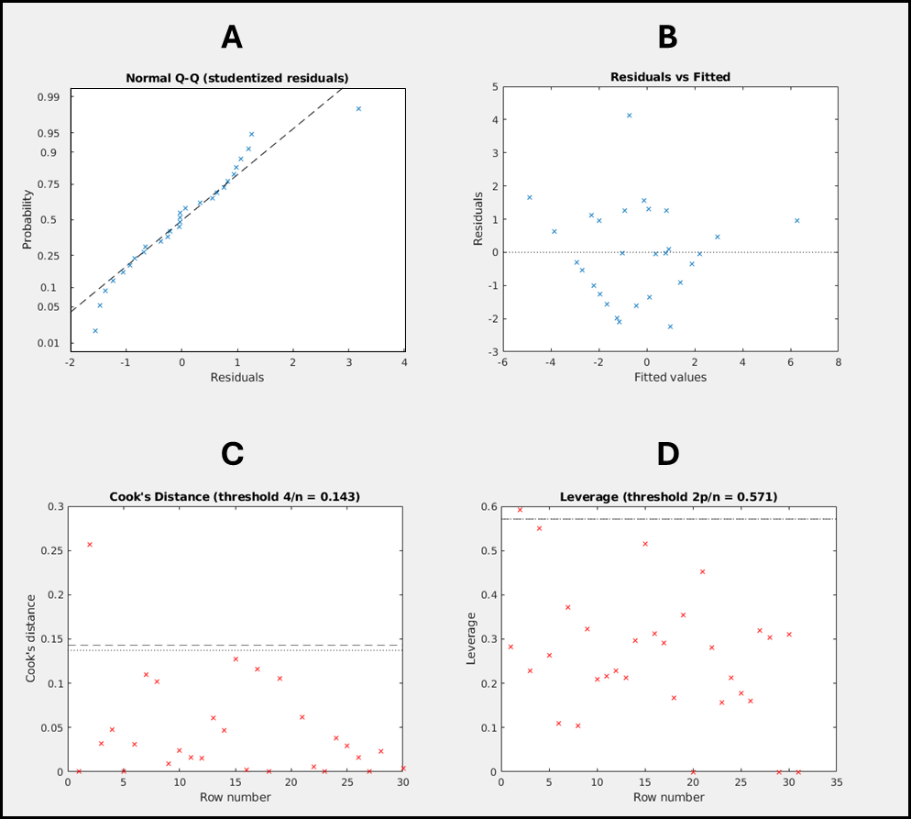

Figure S2. Diagnostic Plots for original maximally-fitting Ordinary Least Squares main effects model.

###### 1.2.4.2. Moderation Analysis

Regression diagnostics for the original OLS interaction model flagged two observations, one with |studentized residuals| > 3 (i.e., 4.95) (Figure S3A) and elevated Cook’s D (0.149, with threshold at 0.143), one with elevated leverage (0.663, with threshold at 0.643) (Figure S3D).

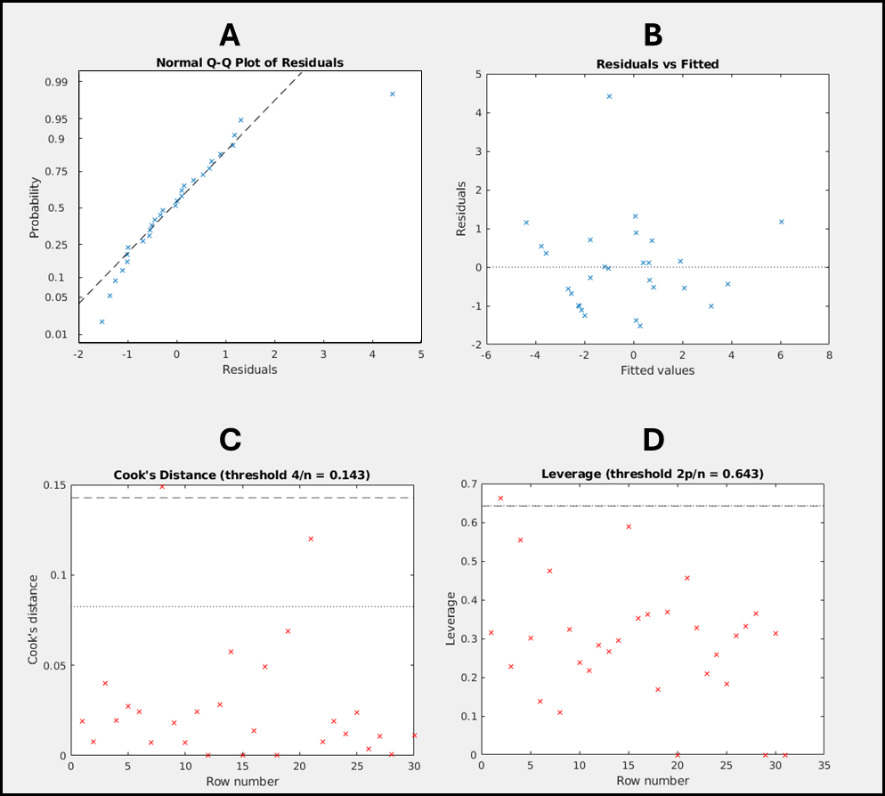

Figure S3. Diagnostic Plots for original maximally-fitting Ordinary Least Squares main effects model.
